# A simple, effective enclosure with disposable coverings for inexpensive containment of aerosolized COVID viruses during tracheal intubation and extubation

**DOI:** 10.1101/2020.11.23.20237255

**Authors:** Luke W. Monroe, Jack S. Johnson, Howard B. Gutstein, John P. Lawrence, Keith Lejeune, Ryan C. Sullivan, Coty N. Jen

## Abstract

**Background:** SARS-CoV-2 (COVID-19) is a severe respiratory virus that can be transmitted through aerosol particles produced by coughing, talking, and breathing. Medical procedures used to treat severe cases such as tracheal intubation, extubation, and tracheal suctioning produce infectious aerosol particles. This presents significant risk for viral exposure of nearby healthcare workers during and following tracheal operations. This study looks at an enclosure to limit medical personnel’s exposure to these particles.

**Methods:** A low-cost plastic enclosure was designed to reduce aerosol spread and viral transmission during intubation and extubation procedures. The enclosure consists of clear polycarbonate for maximum visibility. Large side cutouts provide health care providers with ease of access to the patient. Aerosol particle instruments measured the aerosol containment efficacy after applying various types of plastic coverings to seal the side openings. The use of negative pressure was also tested.

**Results:** The enclosure with 2 layers of plastic coverings sealing the side openings reduced total escaped particle number concentrations (diameter > 0.01 μm) by over 93% at 3 inches away from all openings. Concentration decay experiments indicated that the enclosure without active suction should be left on the patient for 15-20 minutes following a tracheal manipulation to allow sufficient time for >90% of aerosol particles to settle upon interior surfaces. This decreases to 5 minutes when 30 LPM suction is applied.

**Conclusions:** This enclosure is an inexpensive, easily implemented additional layer of protection that can be used to reduce the risk of SARS-CoV-2 aerosol transmission between patients and healthcare workers.

## Background/Introduction

The SARS-CoV-2 pandemic quickly spread globally and has proven persistent in the United States with more than 6.3 million cases and 221,000 deaths (as of Oct. 21^th^, 2020).^1^ The transmission of this virus happens predominantly through three pathways: fomites on contaminated surfaces, contact with large airborne droplets (> 5 µm in particle diameter), and inhalation of smaller aerosol particles (< 5 µm).^2-6^ While droplets and smaller aerosol particles are commonly known to be produced by coughing and sneezing, talking and breathing can also produce infectious aerosols.^2,4-8^ We note that there is no physical distinction between droplets and smaller aerosol particles, but 5 µm in diameter is the conventional boundary used.^2^ Particle size is the main controlling factor governing the transport and suspended lifetime of aerosols.

The transmission vector of the virus through small aerosols is of grave concern due to their long airborne lifetime that allows these particles to travel far distances or disperse through HVAC systems.^2,9-11^ Direct evidence has shown that viable SARS-CoV-2 particles are found in aerosol particles that travel more than 6 ft (1.8 m) from their source. This suggests that the commonly used 6 ft of physical distancing may be ineffective for preventing disease transmission via smaller aerosol particles.^2,11-14^ Aerosols containing the SARS-CoV-2 virus have recently been measured in hospitals.^13,14^ In a clinical setting, modeling routes of transmission suggests that inhalation of aerosol particles, whether near an infectious patient or from dispersed aerosols, is a significant— and perhaps dominant—pathway for exposure and infection of healthcare providers.^15^ This aerosol exposure risk becomes especially pronounced during airway procedures where the patient cannot be masked such as tracheal intubation, extubation, and suctioning. These procedures often result in extensive involuntary patient coughing near healthcare providers.^16,17^

Several approaches have recently been developed to help protect healthcare providers during these procedures, including placing plastic barriers or hoods over the patient in an effort to contain produced aerosols.^16,18,19^ A limitation is that the enclosure must have sizable openings in its side walls to allow the introduction of medical equipment and provide the healthcare worker’s hands access to the patient. These holes have been found to funnel the aerosol that escapes from the enclosure.^16^ The effectiveness of these barriers and feasibility of implementation in a medical setting is variable, with more effective measures such as active ventilation and/or filtration requiring greater infrastructure to operate.^16^ Herein we present aerosol experiments to test the performance of a simple barrier designed for preventing the transmission of larger droplets and smaller aerosol particles during a simulated tracheal manipulation procedure. Consideration of how best to implement this barrier with plastic coverings applied to minimize potential aerosol transmission is experimentally tested and discussed. This enclosure with the use of negative pressure conforms to the FDA guidelines released on August 21^st^, 2020 for such medical devices. Their use without active suction/ventilation is currently not approved by the FDA per these guidelines.^20^

## Materials and Methods

The enclosure is manufactured from polycarbonate by Magee Plastics Company (Warrendale, PA) (**Figure 1**). The sides and top of the material are folded with the seams residing outside to promote easy disinfection of interior surfaces. The dimensions of the enclosure are 16.5 x 18.5 x 24.5 in (41.9 x 47.0 x 62.2 cm) for a total volume of 32.3 gallons (122.5 L). As seen in **Figure 1**, three openings are cut into the sides of the enclosure to allow access to the patient during medical procedures.

**Figure 1:**
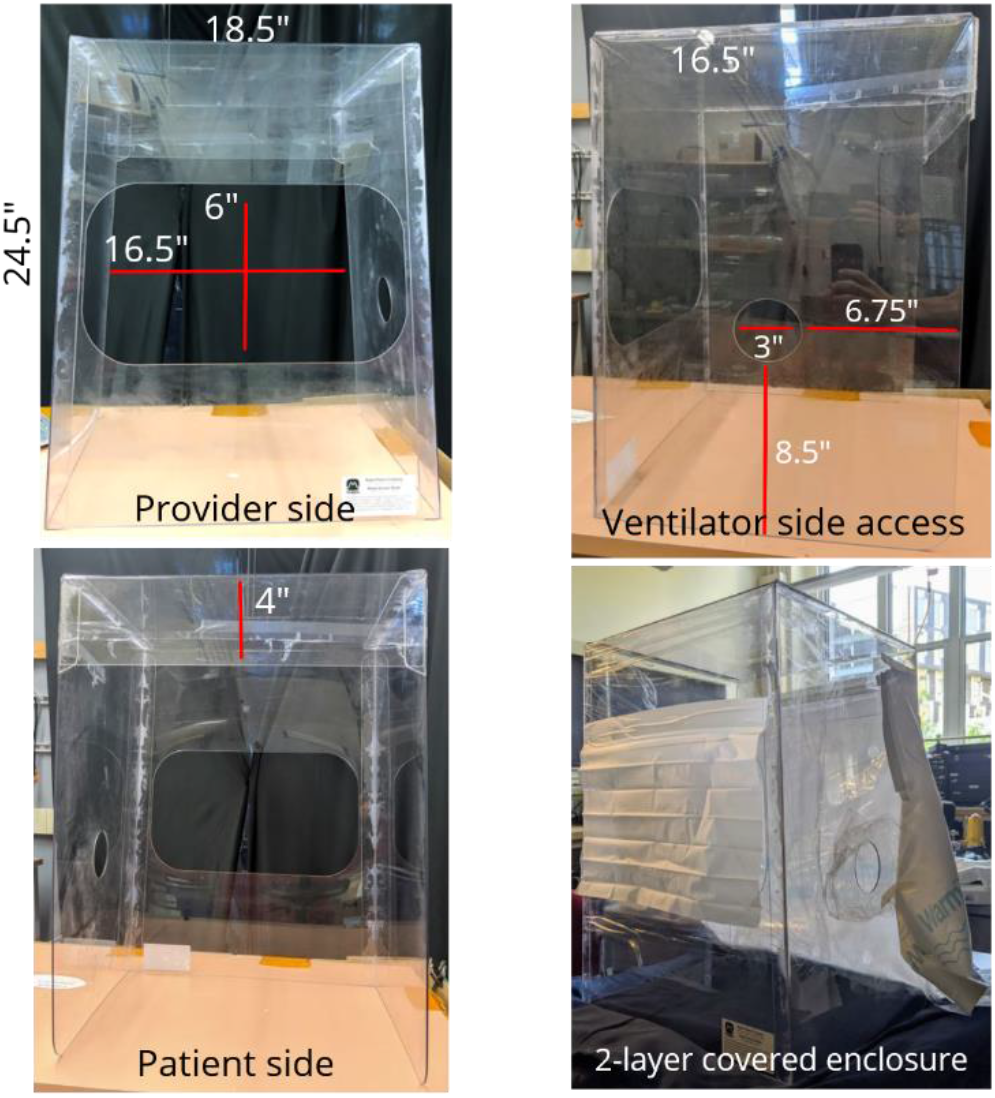
A schematic of the enclosure showing the three cutout openings. One on the cephalad (provider) side to provide access for the healthcare worker’s hands, a second on the caudal (patient) side to allow room for the patient’s torso, and a third on the lateral side to enable ventilator and other tubing access. The provider access cutout is large to maximize mobility.

Various disposable plastic sealing methods for the side and healthcare provider (front) openings were evaluated for their effect on the aerosol containment performance of the enclosure. Sealing methods were chosen based on their ease of procurement and use. Materials tested included Cling Wrap (Glad), Press’n Seal (Glad), Steri 1000 Drape (3M), and 20-inch wide furniture stretch wrap (Goodwrap). Two cross-pattern (+) hand holes were cut into the plastic seal placed over the provider side opening. In some experiments, a Steri-Drape was placed over a layer of either Cling Wrap, Press ‘n Seal, or furniture wrap. In these experiments, the Steri-Drape was not sealed on the bottom and was pushed up by the hands to access the patient. Aerosol instruments measured concentrations outside the enclosure with the sampling lines entering the side hole for internal measurements; the side hole was covered with one layer of the plastic being tested. For all experiments, the large patient-side opening was covered using material from a WarmTouch™ upper body blanket (Covidien) that was composed of an impermeable plastic barrier and a layer of cloth to prevent the movement of the thin plastic covering underneath.

Several nebulizers were used to generate aerosol particles that mimic the diameter and velocity of particles produced by breathing or coughing (size distributions shown in the Supplemental Information, **Figure S1** and **Figure S2**). A Micro Air medical nebulizer (Omron) was used to produce smaller aerosol particles with lower ejection velocity. The medical nebulizer produced two modes in the particle number size distribution centered at 0.18 and 0.6 μm. The large mode is similar to the lower aerosol size range exhaled during normal breathing.^21^ In addition, this nebulizer is a self-contained propellant system similar to human breathing, eliminating positive pressure interference within the enclosure. To produce a size distribution and particle velocity more representative of a cough (i.e., particles with velocities upwards of 32 ft/s)^22^, a Paasche Talon Airbrush was used and produced an aerosol number size distribution with modes at 0.2, 0.5, and 2.3 μm. The Airbrush was chosen based upon studies demonstrating that it generates particles similar in size to evaporated droplet nuclei generated by human coughing (0.74 – 2.12 μm).^23,24^ In all cases the Paasch airbrush was pointed upwards at an angle of 60-75 degrees above horizontal, while the Omron medical nebulizer ejected particles vertically.

Generated particle concentrations were one to two orders of magnitudes higher than those observed by human coughing (**Figure S3**). This was done to simulate ‘worst-case’ scenarios and amplify the aerosol signal that escaped the enclosure, facilitating reliable measurements. Placing the copper sampling lines in areas most prone to particle escape also increased measured aerosol signal.

Negative pressure in the enclosure was produced in some experiments by inserting a vacuum hose (3/8 in) through the side hole. Either a 15 LPM or 30 LPM suction was applied, with a suction canister in line. This setup is typically used in the operating room to remove fluids aspirated from the patient. A small HEPA filter was used in-line. During experiments looking at the impacts of suction, sampling instruments removed 0.3 LPM. When suction was not being considered an experimental variable, the instruments sampled at 3.3 LPM.

In some experiments an investigator’s hands wearing nitrile gloves were inserted through the holes cut in the front covering and then withdrawn to simulate an airway procedure performed on a patient.

All experiments were done with three to five replicates. Error bars and uncertainty values represent the standard deviation. Two-sided statistical analyses (specific tests used cited with experiments below) were conducted using MATLAB 2019a and 2019b, and Microsoft Excel 2016. A p-value less than 0.05 was considered significant.

## Results

Testing for the optimal covering configuration was conducted by spraying a constant stream of particles inside the enclosure for 30 seconds with the Paasch airbrush. Several plastic covering configurations including a control set with no material covering the front or side openings were examined. Comparisons of aerosol concentrations inside versus outside the enclosure were made by taking the ratio of the maximum outside to maximum inside concentration to account for turbulent mixing in the enclosure. The maximum values being compared do not necessarily represent the same time in the run.

The observed number concentrations of 0.01 µm to 10 µm particles inside and 3 in. (7.62 cm) outside the enclosure without plastic covers on the provider side indicated that the enclosure trapped 70 ± 11% of generated particles with 30 ± 10% escaping into the room (**Figure 2**). A single layer covering of furniture wrap increased the aerosol trapping efficiency to 86 ± 6%. The addition of a Steri-Drape over the furniture wrap proved effective; > 97 ± 3% of the aerosols were contained inside the enclosure. Steri-Drape over Press’n Seal was also tested with a trapping efficiency of 99 ± 1%. Adding 15 LPM suction to the Steri-Drape plus furniture wrap set up produced a trapping efficiency of 98 ± 3%. These results indicate that plastic sheet covers over the openings of the enclosure lead to significant reductions in peak aerosol concentrations that escape the enclosure (one-way ANOVA p = <0.001). A Games-Howell test indicated a significant difference between no cover, 1-layer cover, and 2-layer covers. However, no significant difference between the different types of 2-layer configurations, with or without applied suction, was determined.

**Figure 2:**
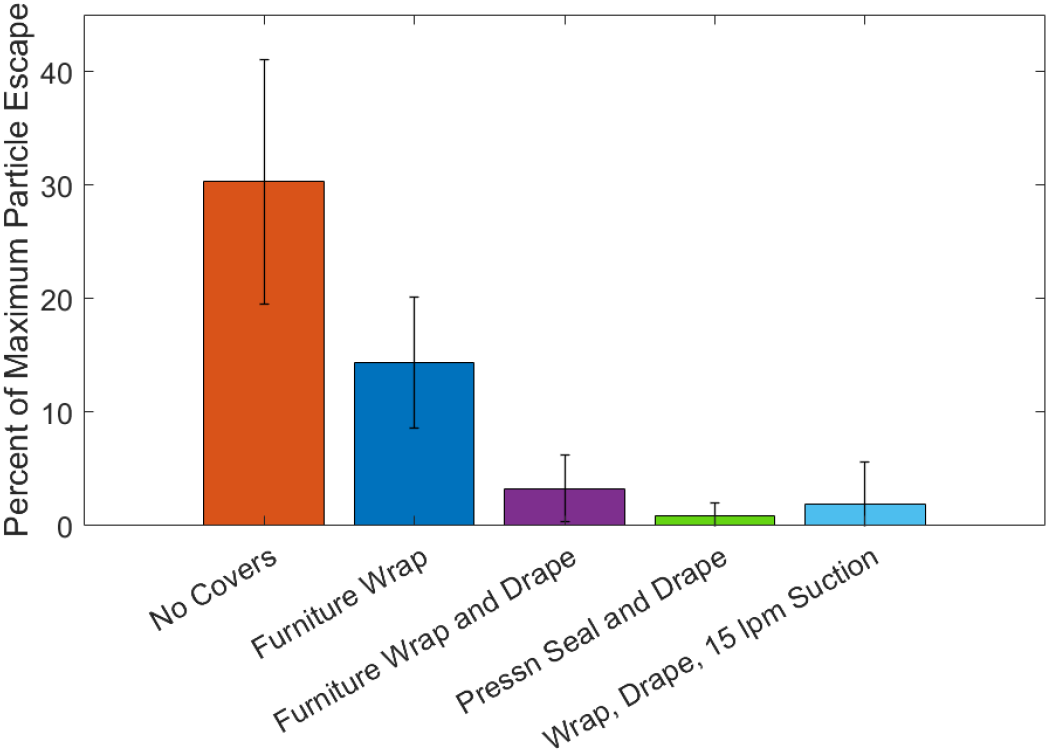
Ratio of the maximum aerosol mass concentration measured inside vs. measured outside the enclosure for particles larger than 0.1 microns when various types of one-layer of plastic covering with or without an added Steri-Drape layer were placed over the front and side holes in the enclosure.

Measurements were conducted at 3, 6, and 12 in. away from the openings to determine at what distance healthcare providers and equipment need to be positioned from the enclosure to avoid exposure from escaping particles. In contrast to the previous experiments, the medical nebulizer was used in the enclosure to generate particles for two minutes with no suction applied. **Figure 3** illustrates the fraction of particles larger than 0.01 μm that escaped from the enclosure with either no coverings (**Fig. 3a**) or Cling Wrap and Steri-Drape (**Fig. 3b**) on the openings. Two-sample t-tests were conducted at each measurement distance between covered and uncovered configurations followed by a Bonferroni correction to determine significance for multiple comparisons. There was a significant reduction in total particle concentration at 3 in. (67 ± 12% no covers versus 7.6 ± 9% with covers, p = <0.001) from the enclosure. After the Bonferroni correction, the results at 6 in. and 12 in. were not statistically significant. This discrepancy is likely a result of the large variability in measured particle concentrations at far distances from the uncovered enclosure. This is evident by the more than doubling in the standard deviation from measurements conducted at 3 in. compared to 6 in. There was not enough particle leakage measured without a cover on the side hole to determine if there was a significant difference between cover and no cover at any distance for the side hole.

**Figure 3:**
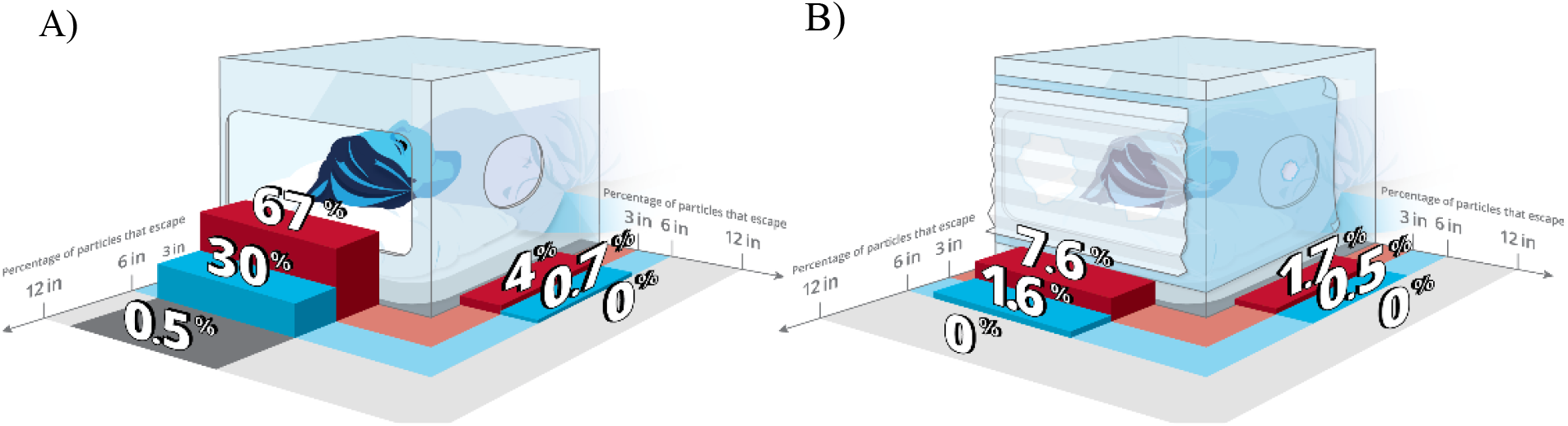
Outside particle concentration compared to inside particle concentration as a function of distance for enclosure (A) without plastic coverings vs. (B) enclosure with dual coverings of furniture wrap and Steri-Drape.

A simulated cough experiment employing the airbrush was used to determine the time required for particles inside the enclosure to settle onto interior surfaces. Air was pulsed through the airbrush in three 1-second bursts to simulate a cough. **Figure 4a** shows the time for particle mass concentrations inside the enclosure to decease by 90% from the peak concentration following the simulated cough event. With no coverings, this was achieved in roughly 4 minutes, likely driven by rapid escape of particles from the enclosure. Adding two layers of covers increased the estimated 90% reduction time inside the enclosure to 14 ±6 min. Particle mass loss of 80% was seen at 5 ± 3 min and 95% loss at 22 ± 12 min. Once suction was applied for the dual-covered enclosure, as shown in **Figure 4b**, the rate of particle loss was observed to greatly increase as suction flow rates were raised. The particle lifetime decreased from 7.1 ± 1 min without suction to 2.0 ± 0.2 min with the application of 30 LPM suction.

**Figure 4:**
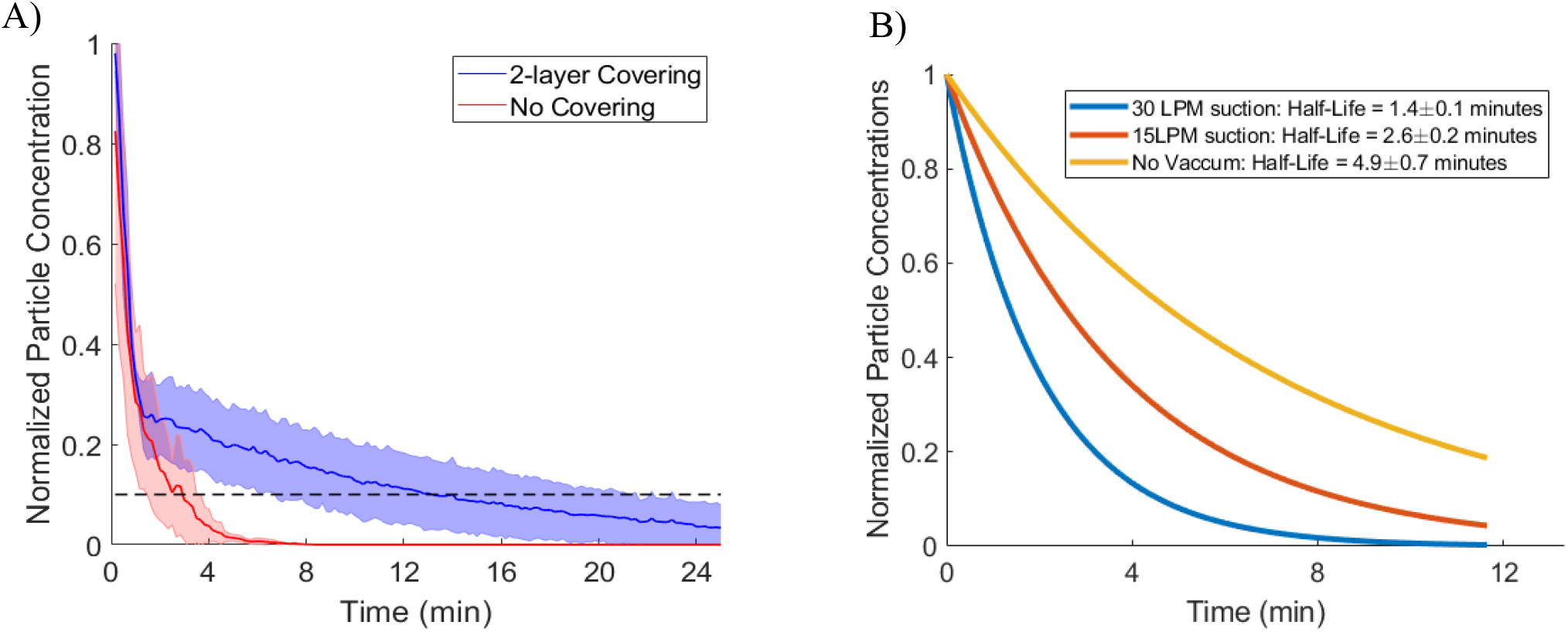
(A) Average decay rate curves for aerosol particle mass concentrations within the enclosure following a particle generation event. The no cover condition kept the provider and ventilator access holes open. The 2-layer cover condition used the furniture wrap and Steri-Drape. Shaded region shows the standard deviation from 3 replicates. The black dashed line indicates 90% particle mass loss. (B) Comparison of average decay rates for aerosol particle mass concentration when suction is applied to the enclosure. All three conditions involve a dual-covered enclosure with furniture wrap and Steri-Drape. The patient side was covered for all experiments.

Typical airway procedures require the provider’s hands to reach into and out of the front opening of the enclosure, potentially jostling the device and leading to increased escape of aerosol particles. Experiments were conducted to determine whether hand movements altered the amount of aerosol particles that escaped the enclosure when covered with furniture wrap and a Steri-Drape. Hands were placed inside the holes cut into the furniture wrap cover either shortly before or shortly after a simulated cough event. Hands were moved while inside the enclosure to simulate a healthcare provider performing a procedure. After 1 minute the hands were removed from the enclosure. The results indicate no statistically significant increase in outside particle concentrations for either approach (**Figure S4**).

Other field testing, such as agitating the enclosure by mildly shaking it, with and without hands in, were conducted. The measurements suggested no significant increase in outside particle concentrations (Data not shown). This indicates that furniture wrap adheres well around the wrists and forearms of the provider. It also indicates that the dual-cover system can withstand general use within a medical setting with the mild bumps and movements.

## Discussion

The measurements show that 1-layer of plastic covering was not the most effective approach to trapping particles inside the enclosure. Adding an additional Steri-Drape on top of the first layer of plastic to create a 2-layer covering was observed to be effective with more than 97 ± 3% of total aerosol particles larger than 0.01 µm contained within the enclosure. However, Glad Cling Wrap in the 2-layer seal configuration proved to be less effective (not shown) compared to furniture wrap. There are two potential reasons for this result. First, the Cling Wrap covering was observed to electrostatically repel the Steri-Drape layer, creating a larger gap between the hand holes in the first layer and the drape that lies over these holes. Second, this material was more fragile than the furniture wrap and tore around the hand holes during use. The Press’n Seal covering appeared to electrostatically attract the Steri-Drape layer, potentially reducing opportunities for particles to escape through the cut hand holes. Both plastics showed reduced tearing at the hand holes during use. The furniture wrap combined with the Steri-Drape proved to be easier to use than the Press’n Seal combined with the Steri-Drape because of its better transparency and self-adhesive properties which enabled it to be secured to the enclosure without extra adhesive. Therefore, we recommend furniture wrap combined with the Steri-Drape to cover openings on the provider side of the enclosure. A single layer of wrap appears to be sufficient for the side hole.

The enclosure with two layers of covers applied over the front opening with no suction applied should remain over the patient for at least 15 minutes after the last particle generation event for 90% particle removal. The addition of suction at 30 LPM and 15 LPM reduces this timescale to 5 and 9 minutes, respectively. Particle losses within the enclosure are driven by several mechanisms depending on particle diameter. Larger particles will impact onto the enclosure if ejected with enough force or gravitationally settle to the bottom. Smaller particles are likely lost by interception with internal surfaces as they circulate within the enclosure, attracted via electrostatic forces, or removed by the suction flow. Particles do not become re-aerosolized once they have deposited on a surface, and thus there is little concern of re-aerosolization if the enclosure is removed from the patient following the settling period. Care should be taken when sterilizing the enclosure after use as fomites would be a concern for transmission. A sheet can be placed over the patient to collect most of the aerosol that settles at the bottom and then either disposed of or handled as contaminated and sterilized.

The experiments conducted with this enclosure captured the dynamics of particles similar to those produced by coughing and breathing. Both the airbrush and medical nebulizer generated particle sizes similar to those produced by passive respiration, speaking, and coughing, as well as even smaller particles (< 0.3 µm).^6,23,24^ The smaller particles are more mobile and difficult to trap than the larger particles produced by coughing. Therefore, these tests can be considered ‘worst-case’ scenarios containing higher concentrations of smaller particles that settle more slowly than would be present in the clinical situation. Nevertheless, we still observe drastic reductions in particle concentrations with the dual layer of plastic coverings without a corresponding increase outside the enclosure. Thus, it is reasonable to conclude that this enclosure will protect against smaller exhaled particles as well as larger cough droplets.

The design and performance of two other intubation enclosures were recently reported.^18,19^ These enclosures utilized a suction device to generate negative pressure and HEPA filters to help prevent the spread of aerosolized particles, with the openings either uncovered or covered with rubber septum. ^18,19^ Phu et al. reported a 99% particle reduction in the 0.5 to 5 µm range, which is slightly higher than the 93-97% reduction reported here without suction. This is likely due to our particle measurements extending down to smaller sizes (to 0.01 µm), because these smaller particles can more easily escape the enclosure.^16^ The passive aspect of the intubation enclosure presented here provides a significant layer of protection to reduce the spread of potentially infectious aerosol without the need for active suction. The benefit of adding active suction to the enclosure is to reduce particle lifetimes in the enclosure.

This study investigates the use of an effective, inexpensive, and easily sterilizable enclosure that utilizes accessible disposable plastic covers. It can be readily implemented to minimize risk of airborne disease transmission to healthcare providers during tracheal operations. Various methods and materials to seal the openings in the enclosure were examined. A two-layer covering on the provider-facing opening consisting of furniture wrap and Steri-Drape on top was found to be the optimal method to reduce aerosol number concentrations escaping the enclosure. The side access hole should be covered with a layer of furniture wrap, and the patient side with a layer of plastic and a layer of heavier fabric on top. Depending on the amount of suction applied, a 5-20-minute waiting period following the last particle generation event is required to allow 90% of particles to settle or be suctioned out before safely removing the enclosure.

## Data Availability

All data can be furnished upon request by reaching out to the corresponding author

## Acknowledgements

Funding for this research was provided by Highmark Health. We thank Peter Freeman of the Department of Statistics and Data Science at Carnegie Mellon University for advice regarding appropriate statistical analysis for this dataset. We also appreciate the help of Tim Kelly for designing Figure 3.

## Figure titles

- Design and suggested cover configuration of enclosure
- Comparison of the effectiveness of different cover configurations in trapping particles within the enclosure
- Reduction of aerosol transmission using the enclosure with and without coverings
- Internal particle decay within the enclosure with and without coverings

