## Supplemental Information for "A simple, effective enclosure with disposable coverings for inexpensive containment of aerosolized COVID viruses during tracheal intubation and extubation"

Table of contents:

Page 3: Aerosol composition and reasoning
Page 3: Instrumentation description and capabilities
Page 4: Figure S1: Small particle size distribution of Omron medical nebulizer and Paasche Talon airbrush
Page 5: Figure S2: Large particle size distribution of Omron medical nebulizer and Paasche Talon airbrush.
Page 6: Figure S3: Total particle concentrations within the box compared to human output
Page 7: Figure S4: results from agitation experiments

**Aerosol particle composition**

Aerosol particles were generated primarily from a 1 mg/mL bulk solution of aqueous ammonium sulfate (Sigma-Aldrich, ACS grade). In addition, particle composition was varied to better mimic organic components in cough droplets using a mixed solution of 2 mg/mL azelaic acid (Acros Organics, 98%) and 10 mg/mL sucrose (Sigma, ACS grade) in deionized water. While most of the relevant aerosol behavior is driven by particle size, composition can potentially influence the electrical properties of the particles (e.g., dielectric constant and polarizability). Hygroscopicity can also influence aerosol behavior. No noticeable difference was observed in particle trapping efficiencies with different particle compositions.

**Instrumentation description and capabilities**

Aerosol measurements were obtained with two condensation particle counters (CPCs, TSI, 3772 and 3775) for total number concentrations, an aerosol monitor (TSI, DustTrak DRX 8533) for aerosol volume/mass size distributions, and an aerodynamic particle sizer (TSI, 3321) for aerosol number size distributions. The CPCs have a 50% diameter cutoff at 10 nm (model 3772) and 4 nm (model 3775). The CPCs therefore measured all aerosol particles present with integration time either 1 or 10 seconds. CPC 3775 was chosen to operate inside the enclosure due to its lower flow rate and ability to measure a higher total number concentration of particles (up to $10^7$ $cm^{-3}$). To determine the particle size distribution for each particle generation method, the CPCs were combined with a differential mobility analyzer to produce a scanning mobility particle sizer (SMPS).

The DustTrak measured particles between 0.1 μm to 15 μm in aerodynamic mass diameter. The APS can nominally detect particles between 0.3 μm to 20 μm, with accurate sizing of particles greater than 0.52 μm in aerodynamic number and mass diameter. The placement of the DustTrak or APS inside the enclosure depended on the test; generally, the DustTrak was used inside due to its ability to handle higher aerosol mass loadings, and the APS was used outside due to its sensitivity at lower aerosol number concentrations.

Aerosol measurements are presented as number ($cm^{-3}$) or mass concentration ($mg/cm^3$) based on the measuring principles of each instrument. The CPCs measured number concentrations, the DustTrak reported volume/mass concentrations, and the APS reported both. Comparisons between inside and outside the enclosure are only made between instruments with similar measurement metrics.

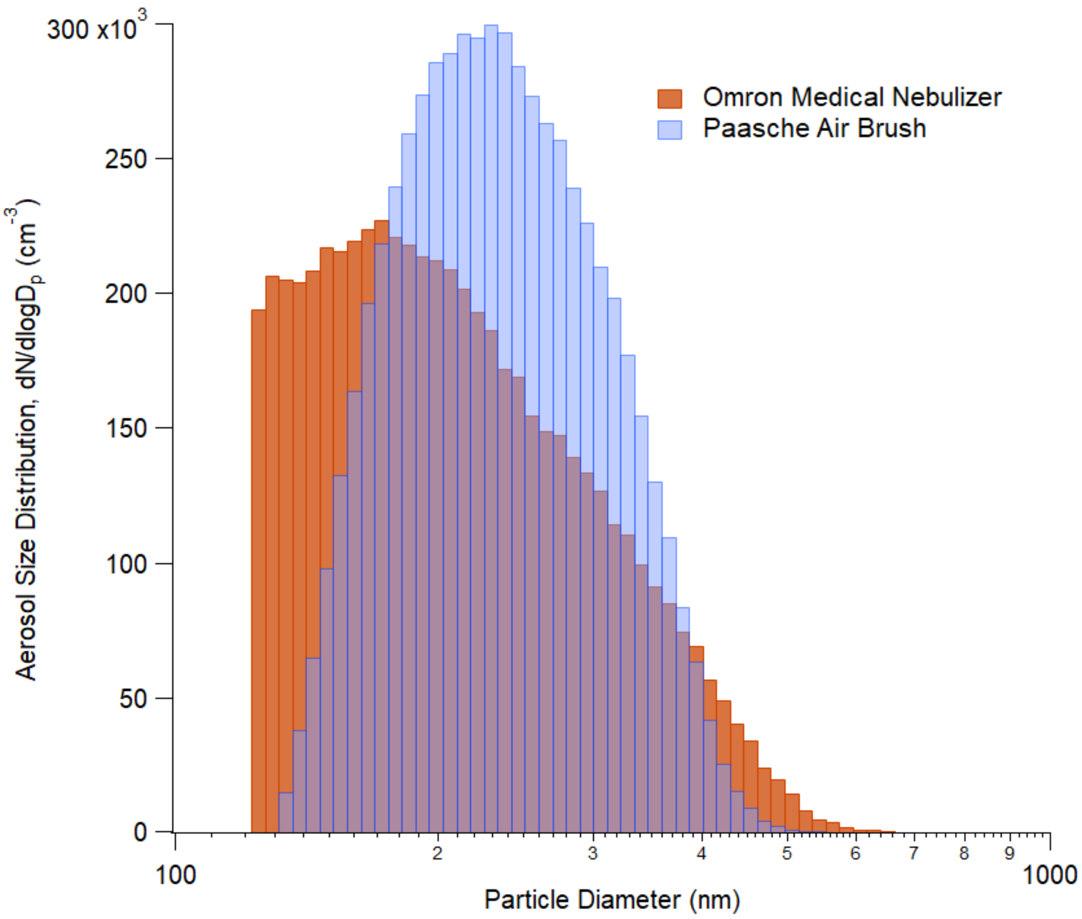


**Figure S1**: Number size distributions of particles between 0.12 µm and 0.6 µm generated by the Paasche airbrush (blue) and Omron medical nebulizer (orange) measured with the scanning mobility particle sizer (SMPS). Particles below 120 nm were not measured to shorten measuring time to limit the impact of the initial rapid particle decay. It is also presumed that viral particles would not be able to be present in particles below this size. The airbrush has a mode at 0.23 µm and the medical nebulizer at 0.17 µm.

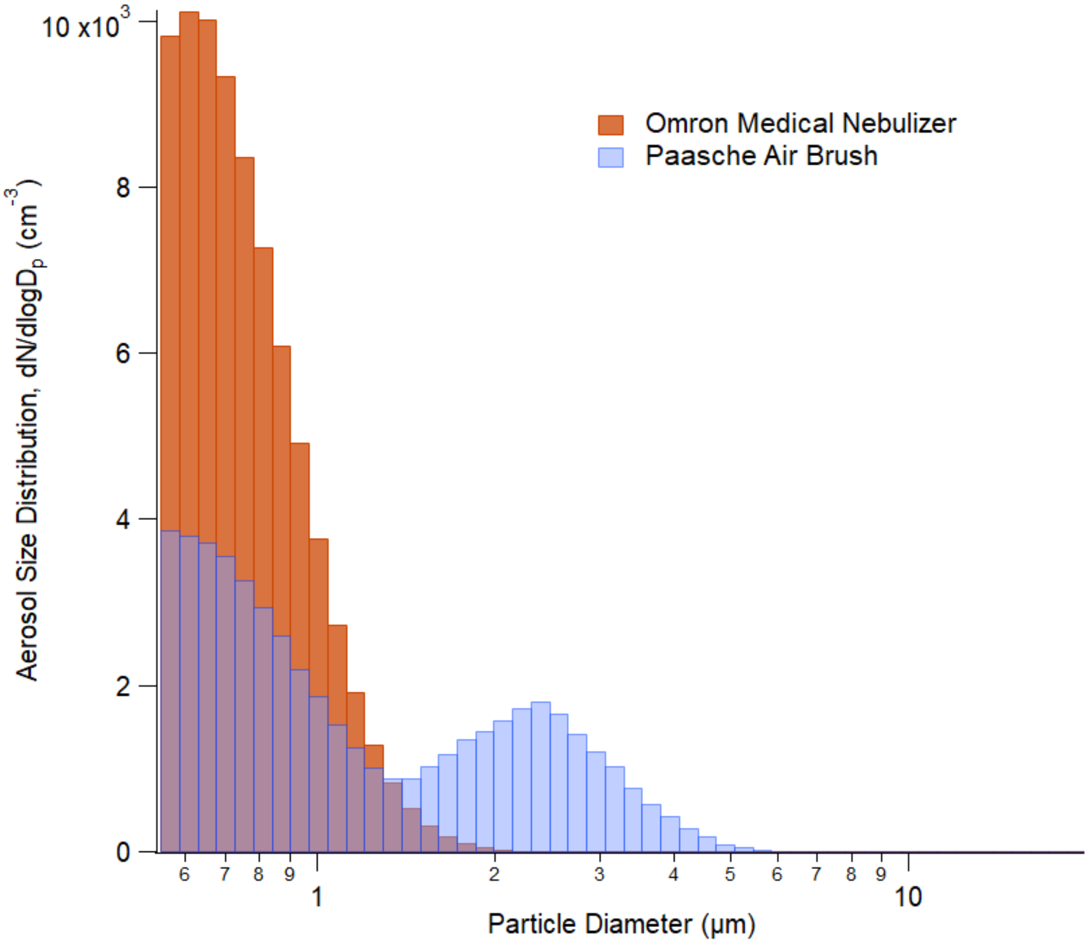


**Figure S2**: Number size distribution measured with the Aerodynamic Particle Sizer (APS) of particles >0.5 µm generated from the Paasche Airbrush (blue) and Omron medical nebulizer (orange). The airbrush has 2 modes, one at 0.6 µm and another at 2.1 µm. The medical nebulizer has one at 0.7 µm.

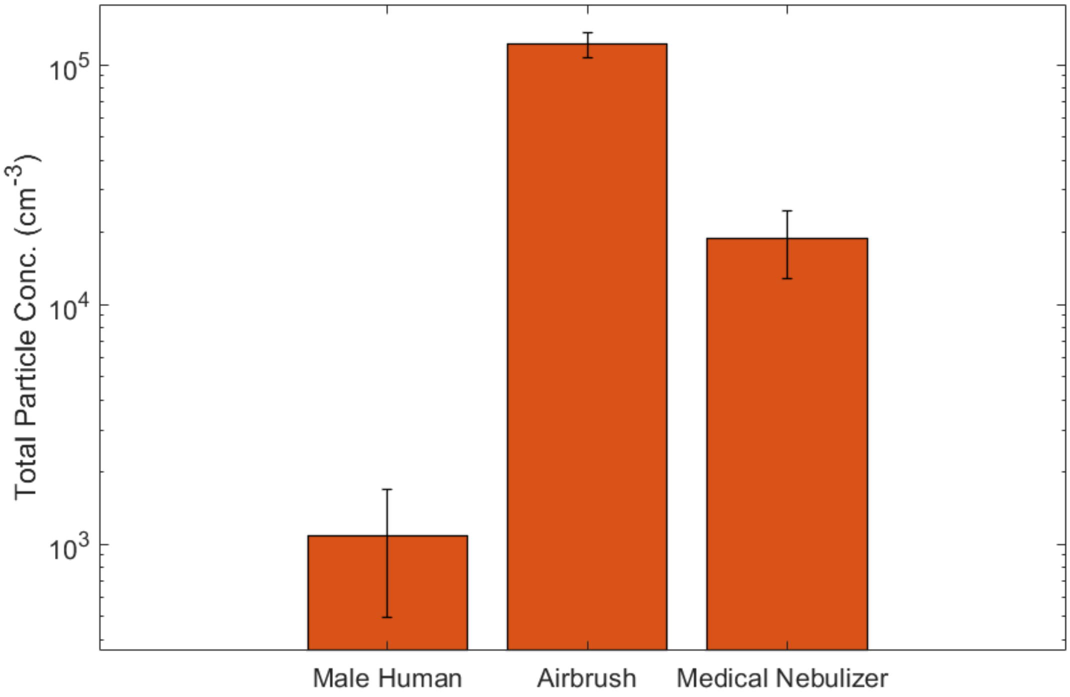


**Figure S3**: Comparison of total particle concentration generated from real and simulated coughs. The average male human cough (Yang et. al, 2007) compared with the simulated Paasche airbrush cough and an extended burst from the Omron medical nebulizer. The medical nebulizer and airbrush means are derived from the peak concentration achieved within the uncovered enclosure.

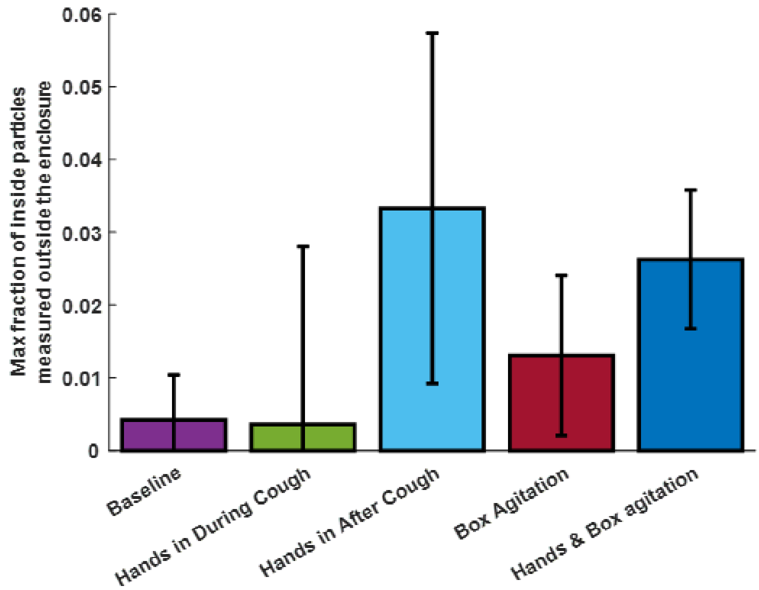


**Figure S4:** Different results of the agitation experiments with error bars representing standard deviation between 3 trials. The 2-layer furniture wrap and Steri-Drape were used for these experiments, with hands covered in nitrile gloves. Baseline is the 2-layer covered enclosure with no hands or jostling. No statistical difference was found with one-way ANOVA. Jostling and inserting/removing hands in the box will not lead to large increases in particle concentrations outside of the box.
